# Depressive symptoms are associated with DNA methylation age acceleration in a cross-sectional analysis of adults over age 50 in the United States

**DOI:** 10.1101/2023.04.24.23289052

**Authors:** Herong Wang, Kelly M. Bakulski, Freida Blostein, Brittany R. Porath, John Dou, Cesar Higgins Tejera, Erin B. Ware

**Affiliations:** School of Public Health, University of Michigan, 1415 Washington Heights, Ann Arbor, MI, 48109, USA; Institute for Social Research, University of Michigan, 426 Thompson St, Ann Arbor, MI, 48104, USA

**Author notes:** **Corresponding author:** Erin B. Ware, 426 Thompson St, Ann Arbor, Michigan 48104.

**Keywords:** Epigenetic clock, DNA methylation age, epigenetic age acceleration, depressive symptoms, epidemiology, older adults

## Abstract

**Background:** Major depressive disorder affects mental well-being and accelerates DNA methylation age, a marker of biological aging. Subclinical depressive symptoms and DNA methylation aging have not been explored.

**Objective:** To assess the cross-sectional association between depressive symptoms and accelerated DNA methylation aging among United States adults over age 50.

**Methods:** We included 3,793 participants from the 2016 wave of the Health and Retirement Study. Depressive symptoms were assessed using the Center for Epidemiologic Studies Depression scale and operationalized as high versus low/no. Blood DNA methylation GrimAge was regressed on chronologic age to obtain acceleration. Multiple linear regression assessed the relationship between high depressive symptoms and GrimAge acceleration, controlling for demographic factors, health behaviors, and cell type proportions. We investigated sex and race/ethnicity stratified associations.

**Results:** Participants were 42% male, 14% had high depressive symptoms, 44% had accelerated GrimAge, and were mean age 70 years. In our fully adjusted model, those with high depressive symptoms had 0.40 (95%CI: 0.06, 0.73) years accelerated GrimAge, compared to those with low/no depressive symptoms. The association between depressive symptoms and GrimAge acceleration was larger in male participants (*P* = 0.04).

**Conclusion:** Higher depressive symptoms were associated with accelerated DNA methylation age among older adults.

## Introduction

Major depressive disorder is characterized by feelings of sadness, hopelessness, and discouragement, or the loss of pleasure or interest in activities for more than 2 weeks^1^. In the United States, approximately 9% of adults aged 65 years and older have a diagnosis of major depressive disorder^2^. Also in the United States, nearly 18% of adults aged 65 and over experience depressive symptoms each year^3^, and older adults experience more depressive symptoms than middle-aged adults^4^. High levels of depressive symptoms are associated with greater mortality risk and increased burden of medical conditions^5,6^. Additionally, depressive symptomatology is associated with poorer social functioning among older people compared to chronic medical conditions such as lung disease, hypertension or diabetes^7^. Thus, understanding the multiple effects of depressive symptoms in vulnerable subgroups of the population is of public health importance. There are existing research documenting disparities in depressive symptoms across sex and among racialized social groups^8^. Specifically in the United States, higher levels of depressive symptoms are reported by women relative to men, as well as Black, Hispanic, and Asian individuals relative to White individuals^8–10^. Therefore, understanding underlying molecular mechanisms between depressive symptoms and the aging process and how sex and race/ethnicity may interact with depressive symptoms is of public health importance.

DNA methylation-based biomarkers are one way to measure biological aging. DNA methylation most commonly occurs at cytosine nucleotides upstream of guanine nucleotides, termed CpG sites^11,12^. Levels of DNA methylation at sets of CpG sites and traits are used to train DNA methylation age estimators^11^. When a person’s estimated DNA methylation age is higher than their chronological age, accelerated DNA methylation age is said to occur. GrimAge is a DNA methylation age estimator trained to predict lifespan^13^. GrimAge predicts all-cause mortality and showed strong association with numerous age-related conditions^14^ and is therefore likely to be sensitive for assessing biological aging related to psychiatric conditions^15^. However, there are few publications investigating the association of DNA methylation age acceleration with depressive symptoms using GrimAge.

DNA methylation age acceleration has been observed in middle-age adults with major depressive disorder. Among 1,130 Dutch participants aged 41 on average, major depressive disorder was associated with 0.64 years accelerated blood DNA methylation age, using a custom estimator^16^. Similarly, among 109 participants aged 40 on average, major depressive disorder was associated 2 years accelerated GrimAge measured in blood^15^. These studies in younger age participants were limited however by small sample size and strict diagnosis of the condition. The role of more prevalent sub-clinical depressive symptoms on biological aging among more vulnerable and diverse samples of older adults is less clear. In light of the insufficient evidence on DNA methylation aging related to depressive symptoms among older adults, we examined the association between depressive symptoms and GrimAge age acceleration in a large and diverse cross-sectional sample (n=3,793) of the United States Health and Retirement Study. We also assessed the effect modification of race/ethnicity and sex, and provided race/ethnicity and sex-specific estimations, given that the lack of evidence on the disparities across sex and among racialized social groups in the association between depressive symptoms and biological aging.

## Methods

### Study Sample

The Health and Retirement Study is an ongoing, nationally-representative, panel study of Americans over 50 years old^17^. The cohort began in 1992 with surveys occurring every two years. The Health and Retirement Study uses a multi-stage area probability design with geographical stratification and oversampling of African-American and Hispanic households. During the 2016 wave, the Health and Retirement Study collected venous blood samples for DNA methylation measures^18^, thus we selected this wave for our cross-sectional analyses. The Health and Retirement Study data is sponsored by the National Institute on Aging (grant number U01AG009740) and is conducted by the University of Michigan. This secondary analysis was approved by the University of Michigan Institutional Review Board (HUM00128220).

Participants provided written informed consent. Data are publicly available at https://hrsdata.isr.umich.edu/data-products.

### Depressive Symptom Measures

Depressive symptoms were assessed by self-report on the modified 8-item Center for Epidemiologic Studies Depression (CES-D) scale^19^. The 8-item version of the CES-D scale is consistent, validated, and reliable in measuring depressive symptoms in older adults^20,21^.

Individual item responses were summed for a final score ranging from 0 to 8. For our primary analyses, we created a binary depressive symptoms variable, indicating whether participants had high (4-8 symptoms) or low/no depressive symptoms (0-3 symptoms), consistent with previous studies^22^. In sensitivity analyses, we considered continuous depressive symptoms and an alternatively dichotomized CES-D score (no symptoms versus 1 or more).

### DNA Methylation Age Acceleration Measures

Venous blood was collected by a trained phlebotomist, using a whole blood EDTA vacutainer blood tube^18^. Samples were shipped overnight to the University of Minnesota Advanced Research and Diagnostic Laboratory and processed within 48 hours of collection^12^.

DNA methylation levels were quantified using the Infinium MethylationEPIC BeadChip (Illumina Inc.) for 4,104 samples randomized across plates by key demographic variables including age, cohort, sex, education, and race/ethnicity. Measures from 86 samples were excluded during quality control assessments^12^. DNA methylation age estimates were calculated and released by the Health and Retirement Study^12^.

Our primary DNA methylation age estimate was GrimAge. GrimAge is trained on health-related and disease-related indicators like sex, education, race, and smoking and is therefore likely to be a stronger and more sensitive predictor for all-causes mortality, age-related health status and biological aging in psychiatric conditions^13–15^. To create the age acceleration values, we regressed the DNA methylation age estimate against chronological age and captured the residuals from this model^13,23^. We modelled these residuals as continuous in linear regressions and for bivariate analyses and we dichotomized the residuals into accelerated aging (residuals > 0) or no age acceleration (residuals ≤ 0).

In sensitivity analyses, we used four additional DNA methylation age estimates calculated and released by the Health and Retirement Study. These additional age estimates included Horvath, Hannum, Levine, and DunedinPoAm38 (MPOA). Horvath and Hannum were selected as DNA methylation estimators of chronologic age. The Horvath age estimator was trained on DNA methylation at 353 CpG sites across 51 different tissues and cell types^24^. The Hannum age estimator was trained on 71 CpGs selected in blood to capture changes in chronological age^25^. Levine and MPOA were selected as measures of phenotypic aging. The Levine age estimator was trained on 513 CpGs to capture the combination of age-related and disease phenotypes^26^. The MPOA algorithm was trained to predict the rate of aging^27^ and we transformed the MPOA rates into years for alignment with the other age estimators. We regressed each DNA methylation age estimate on chronological age and took the residuals from the linear model to obtain age acceleration values. Residuals were modelled as continuous in linear regressions for use in sensitivity analysis.

### Covariate Measures

Sociodemographic information was provided through self-reported baseline interviews including sex (male/female), race/ethnicity (non-Hispanic White, non-Hispanic Black or Other, and Hispanic), childhood socioeconomic status (financial situation before age 16 of well off financially, about average, poor, varied), and education (no degree, GED/high school diploma, and college or more than college). Interviews at the 2016 wave provided age (years), marital status (currently married/not currently married), alcohol use (number of days per week consuming alcohol), cigarette smoking (never, former, current), number of chronic health conditions, and physical activity (levels of energy expended during exercise weighted by Metabolic Equivalent of Task (METs)) (Supplemental Methods). The covariate dataset used for this study was from the RAND Center for the Study of Aging with funding from the National Institute on Aging and the Social Security Administration. Santa Monica, CA produced (July 2022)^28^.

Cell type composition is an important precision variable for DNA methylation analyses^29^. Percentages of granulocytes, monocytes, and lymphocytes were assessed from a complete blood count using a Sysmex XE-21000 instrument^18^ and were released by the Health and Retirement Study with the DNA methylation age estimates^12^.

### Statistical Methods

The included analytic sample was restricted to participants who had complete data for depressive symptoms, DNA methylation age, and all covariates. We visualized sample inclusion and exclusion using a flow chart. We assessed the sample distributions of continuous variables using mean and standard deviation and for categorical variables using count and frequency. To assess the possibility of selection bias, included participants were compared to participants excluded due to missing variables using Wilcoxon rank sum test for continuous variables and Pearson’s chi-squared test for categorical variables. We calculated two-sided *P*-values from these tests at an alpha of 0.05 in all models. In the included sample, we examined the bivariate characteristics similarly based on whether the participant had high or low/no depressive symptoms. We also calculated bivariate characteristics based on dichotomous GrimAge age acceleration.

To check the validity of age-related variables, we examined the pairwise relationships between chronological age, GrimAge, GrimAge age acceleration, and the difference between chronological age and GrimAge. We created scatterplots to visualize these relationships and calculated Pearson correlation coefficients for each pairing.

We constructed three linear regression models to test the association between high depressive symptoms and GrimAge age acceleration. The initial unadjusted model included the binary variable for high depressive symptoms as the exposure and GrimAge age acceleration as the outcome. In the primary adjusted model, we included chronological age, sex, race/ethnicity, highest degree obtained, marital status, percentage of granulocytes, and percentage of monocytes as predictors. We removed percent of lymphocytes from the models to avoid collinearity. In a health behavior adjusted model, we added the number of days per week of alcohol use, smoking status, and number of chronic health conditions to the primary adjusted model.

To evaluate effect modification by race/ethnicity and sex, we applied separate primary and health behavior adjusted models in strata of race/ethnicity and sex and pooled the estimates using generic inverse variance meta-analyses. To examine the significance of the effect modification, we included product terms between depressive symptoms and race/ethnicity and sex in the models. We assessed heterogeneity of associations across strata of race/ethnicity and sex using Cochran’s Q test. If we observed heterogeneity, we used stratification models and reported the estimates in each stratum. Otherwise, we reported the pooled estimates. We performed all statistical analyses using R version 4.0.3. Code to perform all analyses is provided (https://github.com/bakulskilab).

### Sensitivity Analyses

We conducted several sensitivity analyses to assess the robustness of our results. For each sensitivity analysis, we produced an unadjusted model, a primary adjusted model, and a health behavior adjusted model using the same covariates for each model as described above. First, to assess the relationship between the presence of any depressive symptoms and GrimAge age acceleration, we tested the association between an indicator for any depressive symptoms and GrimAge age acceleration. Second, to assess whether depressive symptoms had a linear relationship with accelerated DNA methylation aging, we modelled depressive symptoms as a continuous variable. To determine whether the relationship between depressive symptoms and accelerated DNA methylation aging was specific to the GrimAge estimator, we examined four alternate DNA methylation age estimators including Horvath, Hannum, Levine, and MPOA. Fourth, we examined whether the results differed after further adjustment for physical activity and financial situation before age 16. Finally, we excluded participants older than 85 to assess the robustness of the linear regression model we used to derive GrimAge age acceleration.

## Results

### Descriptive statistics

There were 4,018 eligible participants for our study and the final analytic sample consisted of 3,793 participants with complete data for all analyses (**Figure 1**). Included participants were similar to excluded participants in chronological age, GrimAge, Horvath, Levine, MPOA, and Hannum age (**Supplemental Table 1**); however, included participants were more likely to be non-Hispanic White, married, have a GED/high school diploma as their highest educational attainment, smoke currently, consume alcohol more days per week and have less chronic health condition compared to excluded participants.

**Figure 1.**
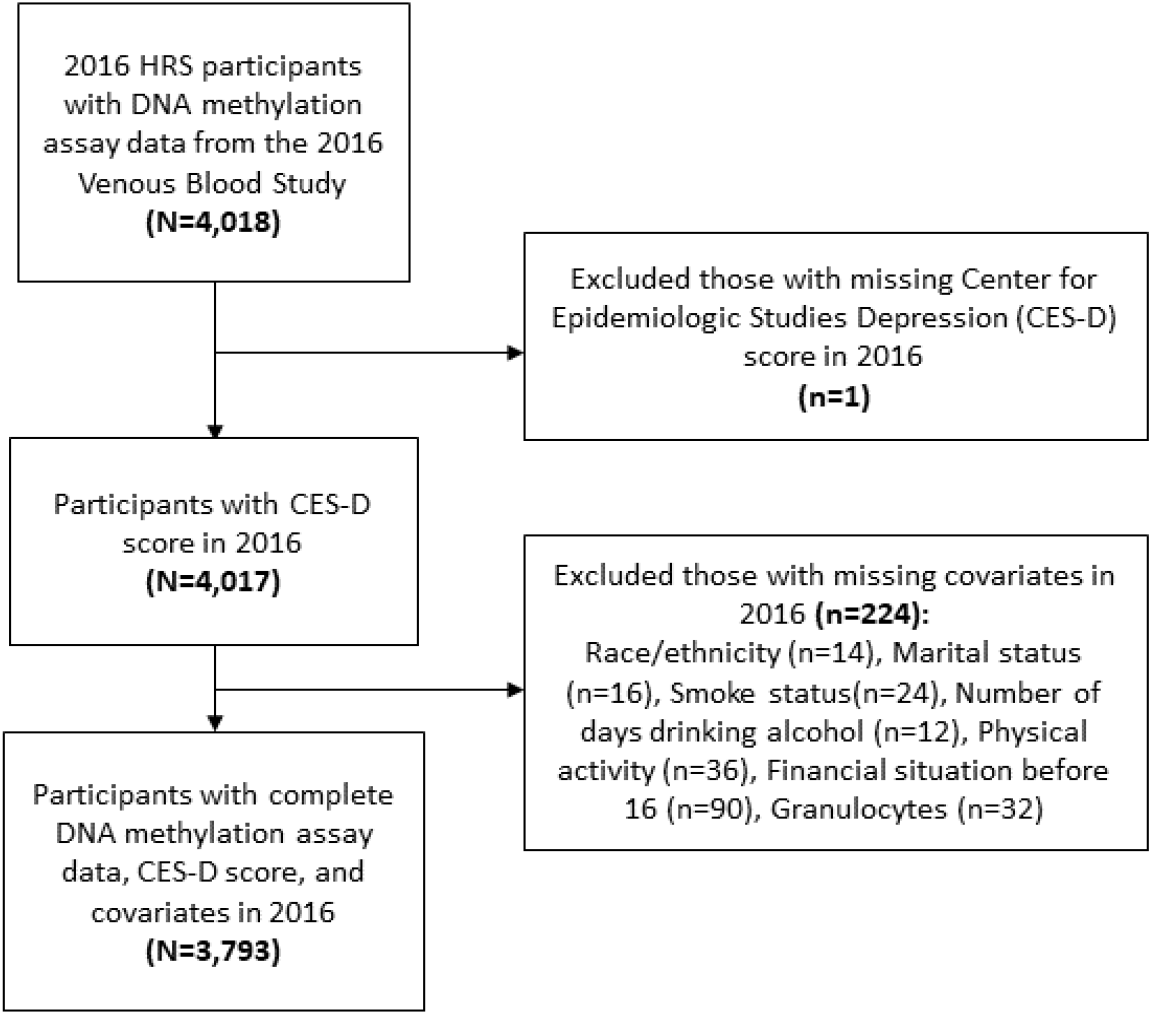
Flow diagram of inclusion and exclusion of participants from the United States Health and Retirement Study 2016 wave who had available DNA methylation data from the 2016 Venous Blood Study. Note: N is the number of observations included. n is the number of observations excluded. CES-D is score on the modified Center for Epidemiologic Studies Depression (CES-D) scale; HRS Health and Retirement Study

Among the analytic sample, 544 (14.4%) of participants had high levels of depressive symptoms based on CES-D score (**Table 1**). Those with high depressive symptoms were significantly more likely to be younger, have higher GrimAge age acceleration, be female, report a race/ethnicity other than non-Hispanic White participants, be single, have more chronic health conditions, be current or former smokers, and have a lower level of physical activity than those with low/no reported depressive symptoms.

**Table 1.** Characteristics of the included analytic sample of the 2016 Health and Retirement Study Venous Blood Study participants with DNA methylation data by level of depressive symptoms and GrimAge age acceleration (n=3,793)

| Characteristic | Overall,<br>N = 3,793 | Low/no depressive<br>symptoms, N = 3,249 | High depressive<br>symptoms, N = 544 | P-value<sup>1</sup> | Non-age<br>accelerated<br>(residuals ≤ 0),<br>N = 2,125 | Age<br>accelerated<br>(residuals > 0),<br>N = 1,668 | P-value<sup>1</sup> |
| --- | --- | --- | --- | --- | --- | --- | --- |
| Chronological age in years, mean(SD) | 70 (10) | 70 (10) | 68 (9) | <0.001 | 70 (9) | 70 (10) | 0.2 |
| GrimAge in years, mean(SD) | 68 (9) | 68 (9) | 68 (8) | 0.2 | 65 (8) | 73 (8) | <0.001 |
| GrimAge acceleration in years,<br>mean(SD) | 0.0 (4.8) | -0.2 (4.7) | 1.1 (5.1) | <0.001 | -3.3 (2.3) | 4.3 (3.5) | <0.001 |
| Has accelerated GrimAge, n(%) |  |  |  | <0.001 |  |  |  |
| Non-age accelerated | 2,125 (56%) | 1,868 (57%) | 257 (47%) |  |  |  |  |
| Age accelerated | 1,668 (44%) | 1,381 (43%) | 287 (53%) |  |  |  |  |
| Horvath age in years, mean(SD) | 66 (10) | 66 (10) | 64 (9) | <0.001 | 65 (9) | 67 (10) | <0.001 |
| Horvath age acceleration in years,<br>mean(SD) | 0.0 (6.4) | 0.0 (6.4) | 0.0 (6.7) | 0.8 | -0.5 (5.7) | 0.6 (7.2) | <0.001 |
| Levine age in years, mean(SD) | 58 (10) | 58 (10) | 57 (10) | 0.029 | 56 (9) | 60 (10) | <0.001 |
| Levine age acceleration in years,<br>mean(SD) | 0 (7) | 0 (7) | 1 (7) | 0.001 | -2 (6) | 2 (7) | <0.001 |
| MPOA age per chronological year,<br>mean(SD) | 1.08 (0.09) | 1.07 (0.09) | 1.09 (0.09) | <0.001 | 1.03 (0.07) | 1.13 (0.09) | <0.001 |
| MPOA age acceleration in years,<br>mean(SD) | 0.0 (6) | -0.2 (6.4) | 1.2 (6.4) | <0.001 | -3 (5) | 4 (6) | <0.001 |
| Hannum age in years, mean(SD) | 55 (9) | 55 (9) | 53 (9) | <0.001 | 54 (9) | 56 (10) | <0.001 |
| Hannum age acceleration in years,<br>mean(SD) | 0.0 (5.2) | -0.1 (5.2) | 0.2 (5.5) | 0.3 | -0.9 (4.8) | 1.1 (5.5) | <0.001 |
| Depressive symptoms n(%) |  |  |  |  |  |  | <0.001 |
| Low/no depressive symptoms | 3,249 (86%) |  |  |  | 1,868 (88%) | 1,381 (83%) |  |
| High depressive symptoms (CES-D score $\geq 4$) | 544 (14%) | | | | 257 (12%) | 287 (17%) | |
| **CES-D score<sup>2</sup>, mean (SD)** | 1.40 (1.96) |  |  |  | 1.24 (1.86) | 1.61 (2.07) | <0.001 |
| **Sex, n(%)** |  |  |  | <0.001 |  |  | <0.001 |
| Male | 1,578 (42%) | 1,408 (43%) | 170 (31%) |  | 638 (30%) | 940 (56%) |  |
| Female | 2,215 (58%) | 1,841 (57%) | 374 (69%) |  | 1,487 (70%) | 728 (44%) |  |
| **Race/ethnicity, n(%)** |  |  |  | <0.001 |  |  | <0.001 |
| Non-Hispanic White | 2,539 (67%) | 2,246 (69%) | 293 (54%) |  | 1,487 (70%) | 1,052 (63%) |  |
| Non-Hispanic Black or Other | 747 (20%) | 610 (19%) | 137 (25%) |  | 352 (17%) | 395 (24%) |  |
| Hispanic | 507 (13%) | 393 (12%) | 114 (21%) |  | 286 (13%) | 221 (13%) |  |
| **Marital status, n(%)** |  |  |  | <0.001 |  |  | <0.001 |
| Married | 2,185 (58%) | 1,968 (61%) | 217 (40%) |  | 1,293 (61%) | 892 (53%) |  |
| Single | 1,608 (42%) | 1,281 (39%) | 327 (60%) |  | 832 (39%) | 776 (47%) |  |
| **Highest level of education, n(%)** |  |  |  | <0.001 |  |  | <0.001 |
| No degree | 628 (17%) | 487 (15%) | 141 (26%) |  | 292 (14%) | 336 (20%) |  |
| GED/high school diploma | 1,987 (52%) | 1,711 (53%) | 276 (51%) |  | 1,041 (49%) | 946 (57%) |  |
| College and more than college | 1,178 (31%) | 1,051 (32%) | 127 (23%) |  | 792 (37%) | 386 (23%) |  |
| **Number of chronic health conditions out of 8<sup>3</sup>, mean(SD)** | 2.43 (1.53) | 2.26 (1.45) | 3.45 (1.60) | <0.001 | 2.18 (1.45) | 2.75 (1.57) | <0.001 |
| **Cigarette smoking status, n(%)** |  |  |  | <0.001 |  |  | <0.001 |
| Never | 1,670 (44%) | 1,452 (45%) | 218 (40%) |  | 1,278 (60%) | 392 (24%) |  |
| Former | 1,685 (44%) | 1,459 (45%) | 226 (42%) |  | 816 (38%) | 869 (52%) |  |
| Current | 438 (12%) | 338 (10%) | 100 (18%) |  | 31 (1.5%) | 407 (24%) |  |
| **Number of days per week consuming alcohol, mean (SD)** | 1.21 (2.03) | 1.25 (2.05) | 0.95 (1.87) | <0.001 | 1.19 (2.01) | 1.22 (2.05) | >0.9 |
| **Physical activity level<sup>4</sup>, mean (SD)** | 23 (15) | 24 (15) | 16 (14) | <0.001 | 25 (15) | 20 (15) | <0.001 |
| **Financial situation before age 16, n (%)** |  |  |  | <0.001 |  |  | 0.088 |
| Well off | 300 (7.9%) | 260 (8.0%) | 40 (7.4%) |  | 174 (8.2%) | 126 (7.6%) |  |
| Average | 2,393 (63%) | 2,089 (64%) | 304 (56%) |  | 1,365 (64%) | 1,028 (62%) |  |
| Poor | 1,100 (29%) | 900 (28%) | 200 (37%) |  | 586 (28%) | 514 (31%) |  |
| **Granulocytes percent, mean (SD)** | 62 (10) | 62 (9) | 61 (10) | 0.3 | 60 (9) | 64 (10) | <0.001 |
| **Monocytes percent, mean (SD)** | 8.53 (2.44) | 8.59 (2.44) | 8.20 (2.37) | <0.001 | 8.51 (2.34) | 8.56 (2.55) | 0.9 |
| **Lymphocytes percent, mean (SD)** | 30 (9) | 30 (9) | 31 (10) | 0.047 | 32 (9) | 28 (9) | <0.001 |

<sup>1</sup>P-value was calculated using the Wilcoxon rank sum test for continuous variables; P-value was calculated using Pearson's Chi-squared test for categorical variables
<sup>2</sup>Center for Epidemiology Studies Depression Scale
<sup>3</sup>Number of chronic health condition out of high blood pressure, diabetes, cancer, lung disease, heart disease, stroke, psychiatric problems, and arthritis
<sup>4</sup>Physical activity level was calculated by summing up frequencies of each physical activity intensity multiplied by matched average Metabolic Equivalent of Task (METs)

We present the distribution of GrimAge age acceleration as were all other age acceleration variables in **Supplemental Figure 1**. GrimAge and chronological age were positively correlated in the analytic sample (Pearson rho = 0.83). We verified the effect of chronologic age was successfully removed from our measure of GrimAge acceleration by plotting GrimAge acceleration against chronological age (Pearson rho=0.002, **Supplemental Figure 2**).

In the analytic sample, 44% of participants had GrimAge acceleration greater than 0 (**Table 1**). Prevalence of high depressive symptoms was higher among those with accelerated GrimAge (17%) relative to those without accelerated GrimAge (12%) (*P* < 0.01). Participants with accelerated GrimAge were more likely to be male, non-Hispanic Black or Other race/ethnicity, and have lower educational attainment. These participants also had more chronic health conditions and higher levels of drinking compared to those without accelerated GrimAge.

### Multivariable associations between depressive symptoms and DNA methylation age acceleration

We performed linear regression models to test whether high depressive symptoms are associated with GrimAge acceleration (**Table 2**). In the unadjusted model, those with high depressive symptoms had 1.29 (95% CI: 0.86, 1.72) years higher GrimAge acceleration on average compared to those with low/no depressive symptoms. The strength of the association was attenuated after controlling for demographic factors and cell-type composition (*β* = 1.15 years, 95% CI: 0.77, 1.54). We observed further attenuation after additional adjustment for health behaviors (*β*= 0.40 years, 95% CI: 0.06, 0.73). Model fit based on adjusted R^2^ value and likelihood ratio test both improved significantly after adjustment for these factors.

**Table 2.** Linear regression models predicting GrimAge acceleration based on presence of high depressive symptoms in the analytic sample of the 2016 Health and Retirement Study Venous Blood Study participants with DNA methylation data (n=3,792)

|  | **Coefficient for high depressive symptoms (CES-D score $\geq 4$)** | **95% confidence interval** | **P-value** | **Model adjusted R<sup>2</sup>** |
| --- | --- | --- | --- | --- |
| Unadjusted | 1.29 | 0.86, 1.72 | <0.001 | 0.01 |
| Primary model<sup>1</sup> | 1.15 | 0.77, 1.54 | <0.001 | 0.25 |
| Health behavior model<sup>2</sup> | 0.40 | 0.06, 0.73 | 0.02 | 0.47 |
| +Financial situation before 16<sup>3</sup> | 0.40 | 0.06, 0.73 | 0.02 | 0.47 |
| + Financial situation before 16 and physical activity<sup>3</sup> | 0.26 | -0.08, 0.59 | 0.14 | 0.48 |

<sup>1</sup>The primary model is adjusted for age, sex, race, highest degree obtained, marital status, percentage of granulocytes, and percentage of monocytes.
<sup>2</sup>The health behavior model is adjusted for all covariates in the primary model and number of chronic health conditions, number of days per week with alcohol consumption, and smoking status.
<sup>3</sup>Sensitivity model is adjusted for all covariates in the health behavior model.

### Stratified sample regression results

After adjusting for sociodemographic covariates and cell type composition in primary model, the association between depressive symptoms and GrimAge acceleration was larger in male participants (*P* = 0.04) (**Supplemental Table 5**). While we found significant effect modification of the association between depressive symptoms and GrimAge age acceleration by sex in primary models, we did not observe effect modification by sex when adding the health behaviour covariates. Nor did we observe effect modification of the relationship between depressive symptoms and GrimAge age acceleration by self-reported race/ethnicity. We show these results in **Figure 2**. Therefore, we do not report stratified models or explore them further in sensitivity models.

**Figure 2.**
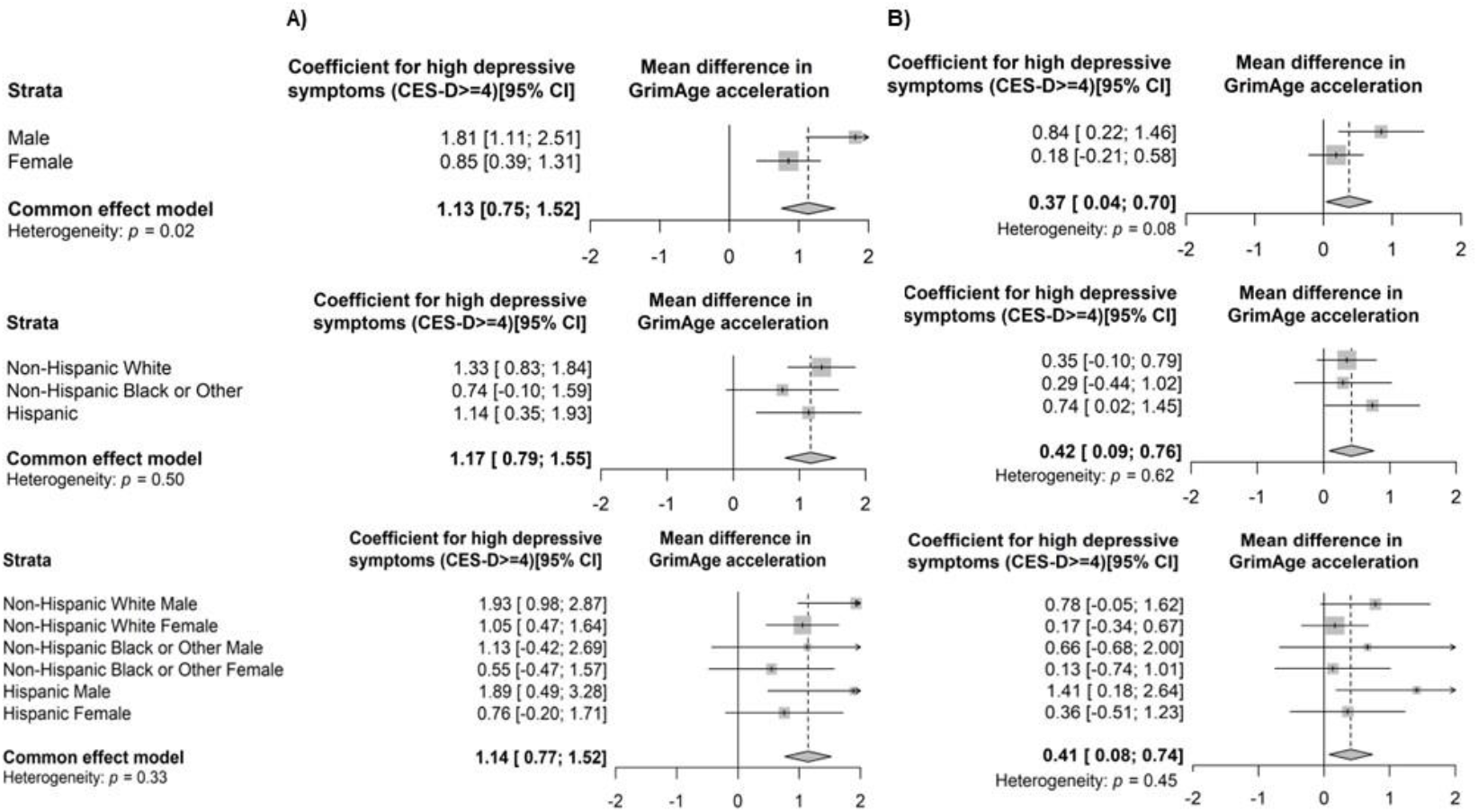
Forest plots for the meta-analysis of race/ethnicity and sex specific regression models of the association between high depressive symptoms (>4) and GrimAge age acceleration among the analytic sample of the 2106 Health and Retirement Study Venous Blood Study participants with DNA methylation data (n = 3,793) Abbreviation: CI, confidence interval. The heterogeneity test and *P*-value were performed using Cochran’s Q test ^A^ The forest plot for race/ethnicity and sex stratified effect estimates and 95% confidence interval in primary models adjusted for age, gender, highest degree obtained, marital status, percentage of granulocytes, and percentage of monocytes ^B^ The forest plot for race/ethnicity and sex stratified estimates and 95% confidence interval in health behavior models adjusted for all covariates in the primary model and number of chronic health conditions, number of days per week with alcohol consumption, and smoking status

### Sensitivity analysis results

In sensitivity analyses examining the parameterization of depressive symptoms, the association between the depressive symptom characterization and GrimAge acceleration remained significant for both 1) any versus no depressive symptoms and for 2) the continuous depressive symptoms measure (**Supplemental Table 2**). When examining other DNA methylation age estimators, the presence of high depressive symptoms was not associated with DNA methylation age acceleration for the Horvath or Hannum clock in any models. The presence of high depressive symptoms was significantly associated with Levine (*β* = 0.73 years, 95% CI: 0.12, 1.34) and MPOA (*β* = 0.93 years, 95% CI: 0.39, 1.47) acceleration in primary models but not in health behavior adjusted models (**Supplemental Table 3**). Smaller and nonsignificant association between high level of depressive symptoms and GrimAge acceleration was observed (0.26, 95% CI: -0.08, 0.59) after further adjustment for physical activity and financial situation before age 16 (**Table 2**) The association between depressive symptoms and GrimAge acceleration in the health behavior model was robust to the exclusion of participants older than 85 (**Supplemental Table 4**).

## Discussion

This cross-sectional study examined the relationship between depressive symptoms and accelerated DNA methylation age among a large, diverse sample of Americans over 50 years old. Among 3,793 participants, the presence of high depressive symptoms was associated with accelerated DNA methylation aging, measured using GrimAge, after adjusting for demographic factors, health status, and health behaviors. We found the association between depressive symptoms and accelerated DNA methylation aging to be robust to different parameterization of depressive symptoms and to other second-generation DNA methylation age estimates, like Levine and MPOA. We ultimately found no effect modification by sex or self-reported race/ethnicity in the association between depressive symptoms and GrimAge acceleration in the fully adjusted model.

We investigated depressive symptoms as an exposure to capture the effects of potential preclinical indicators of depressive disorder on DNA methylation aging. Depressive symptoms affect a larger share of the population than clinically diagnostic forms of major depressive disorder. Previous studies have generally focused on major depressive disorder. Existing studies have found varying effect sizes for the relationship between major depressive disorder and accelerated DNA methylation age. These effects range from an average DNA methylation age estimate of 0.64 years higher for those with major depressive disorder than controls^16^, to around a 2 year increased median GrimAge acceleration for those with major depressive disorder compared to controls^15^. These samples were from middle-aged participants with a mean age around 40 years^15,16^. Additionally, postmortem brain tissue for those with major depressive disorder has shown an average increase of 1.11 years for the Horvath DNA methylation age acceleration compared to controls^16^. Our observation of an increase of 0.40 years in GrimAge DNA methylation age acceleration for those with high depressive symptoms is in line with these results. Major depressive disorder involves an increased burden of depressive symptoms for an extended period that interfere with daily life and represent a more severe phenotype than our dichotomy of depressive symptom counts. In sensitivity analyses, we also observed a significant, linear association between depressive symptoms treated as a continuous variable and accelerated GrimAge DNA methylation where each additional depressive symptom increased the GrimAge age acceleration by 0.08 years. We again would expect a decreased effect size for an increase of individual depressive symptoms compared to a clinical diagnosis of major depressive disorder, especially in older adults where there may be a greater accumulation of environmental effects on DNA methylation age acceleration, which may reduce the individual effect of depressive symptoms.

In our sensitivity analyses, a continuous measure of depressive symptoms (CES-D score) was associated with accelerated GrimAge, after full adjustment. Since the majority of studies in this area focus on major depressive disorder and/or use clocks other than GrimAge, this is a novel finding. These results point to a small dose-response relationship between depressive symptoms and GrimAge acceleration. We performed a second sensitivity analysis to assess the cut-point we selected for determining the presence of depressive symptoms. Any depressive symptoms remained associated with accelerated GrimAge, with an effect size only slightly smaller when compared to the effect size of high depressive symptoms (≥4) on accelerated GrimAge. This suggests that having any level of depressive symptoms may be associated with GrimAge acceleration and that depressive symptoms may be association linearly with increased DNA methylation age acceleration.

Consistent with previous literature, we found that depressive symptoms were associated with “second generation” DNA methylation aging measures, whereas we found no association for the “first generation” DNA methylation aging measures. These second-generation measures of phenotypic aging such as GrimAge, Levine and MPOA, are trained on health indicators such as sex, education, race, and smoking and more highly associated with age-related disease, and thus is likely to be sensitive to psychiatric disorders while measures of chronological aging such as Horvath and Hannum are not. In prior studies, GrimAge was a strong predictor of time-to multiple outcomes, including death, coronary heart disease, and cancer^13^. When examining depressive symptomology among a cohort of 724 Danish monozygotic twins, depressive symptomology score was not associated with Horvath age acceleration, after adjustment for chronological age^30^. These results are consistent with the sensitivity analysis we conducted using the Horvath clock, where we did not observe an association between depressive symptoms and accelerated DNA methylation aging.

The biological mechanism through which depressive symptoms may lead to accelerated DNA methylation age is unknown. Chronic diseases such as major depressive disorder could be prolonged stress exposures, and researchers found chronic stress have multiple physical and epigenetic effects throughout the body^31,32^. Increases in depressive symptoms could also result in behavioral changes, such as smoking, less physical activity, and dietary changes^33–36^ which are risk factors for epigenetic effects^37–39^ and likely to mediate depressive symptoms in DNA methylation aging. When controlling for some such potential health behaviors, we did observe an attenuation of the association, but a significant statistical association remained. Prolonged exposure and increased severity of depressive symptoms may produce chronic stress throughout the body that affects DNA methylation and other cellular characteristics.

This study has several limitations that should be considered when interpreting these results. The cross-sectional nature of this analysis prevents us from establishing that depressive symptoms precede DNA methylation age acceleration, and future work in longitudinal settings is necessary. In addition, more research is needed to understand how the cumulative experience of depressive symptoms across an individual’s lifetime may accelerate DNA methylation aging.

Tissue specificity of DNA methylation signatures may be a concern in considering these results. We used DNA methylation age measured from whole blood, though the brain is the preferred target tissue for these measures when looking at depressive symptoms^40^. However, blood DNA methylation may correlate with brain DNA methylation and whole blood is useful in cases where brain tissue would not be available, such as when using large, population-based studies of living participants^41^. Future research into this area could focus on longitudinal designs while controlling for additional confounders and potentially investigating DNA methylation in disease-specific tissue. Finally, for the purposes of this analysis, we did not use sampling weights, so our results are only generalizable to people over 50 years old living in the United States with similar demographic characteristics to the sample. Future research should investigate whether this association is robust across different scales of measuring depressive symptoms and in other populations.

We were able to interrogate a large number of participants in this study due to the availability of our measures of interest in a population-representative study. Additionally, we were able to adjust for cell type proportions in our analysis, an important precision variable in measuring DNA methylation aging. Our focus on depressive symptoms rather than major depressive disorder diagnosis makes these results more generalizable to those experiencing varying levels and severity of depressive symptoms. We also provided race/ethnicity and sex specific estimates, providing insight on race/ethnicity and sex differences in the association between depressive symptoms and DNA methylation age acceleration. Our use of the GrimAge clock is another strength of this study, since GrimAge predicts lifespan^13^. People with high levels of depressive symptoms typically have higher mortality and more comorbidities, so the GrimAge DNA methylation age estimate is particularly relevant in estimating the potential effect of depressive symptoms^5,6^. Due to these strengths, our study can provide some continuing evidence for an association between depressive symptoms and accelerated DNA methylation age.

In summary, in this cross-sectional analysis we observed high depressive symptoms were associated with accelerated GrimAge among participants in the Health and Retirement Study.

Future studies could try to replicate these findings in other populations and establish a temporal relationship between experiencing depressive symptoms and GrimAge acceleration for the purpose of understanding the effects of depressive symptoms over the life course. Depressive symptoms have likely increased since the beginning of the COVID-19 pandemic, with one study indicating a prevalence of 8.5% among adults in the United States pre-pandemic and a prevalence of 27.8% in April 2020^42^. As depressive symptoms increase in the population, it may be important to study their long-term impact on epigenetic effects. Identifying risk groups, like those with high depressive symptoms, for biological correlates that may lead to earlier mortality could aid in creating intervention targets and highlight the impact of improving mental health management to help reduce early mortality.

## Data Availability

All data produced in the present study are available upon reasonable request to the authors

